# Modeling risk of infectious diseases: a case of Coronavirus outbreak in four countries

**DOI:** 10.1101/2020.04.01.20049973

**Authors:** Md. Mazharul Islam, Md. Monirul Islam, Md. Jamal Hossain, Faroque Ahmed

## Abstract

**Background:** The novel coronavirus (2019-nCOV) outbreak has been a serious concern around the globe. Since people are in tremor due to the massive spread of Coronavirus in the major parts of the world, it requires to predict the risk of this infectious disease. In this situation, we develop a model to measure the risk of infectious disease and predict the risk of 2019-nCOV transmission by using data of four countries—US, Australia, Canada and China.

**Methods:** The model underlies that higher the population density, higher the risk of transmission of infectious disease from human to human. Besides, population size, case identification rate and travel of infected passengers in different regions are also incorporated in this model.

**Results:** According to the calculated risk index, our study identifies New York State in United States (US) to be the most vulnerable area affected by the novel Coronavirus. Besides, other areas (province/state/territory) such as Hubei (China, 2^nd^), Massachusetts (US, 3^rd^), District of Columbia (US, 4^th^), New Jersey (US, 5^th^), Quebec (Canada, 20^th^), Australian Capital Territory (Australia, 29^th^) are also found as the most risky areas in US, China, Australia and Canada.

**Conclusion:** The study suggests avoiding any kind of mass gathering, maintaining recommended physical distances and restricting inbound and outbound flights of highly risk prone areas for tackling 2019-nCOV transmission.

## 1. Introduction

Novel Coronavirus, Middle East Respiratory Syndrome-Coronavirus (MERS-CoV) related to Coronavirus that caused Severe Acute Respiratory Syndrome (SARS) epidemic 10 years ago, was first detected in Saudi Arabia and it has been spreading in other countries since 2012[1]. In December 2019, China noticed few pneumonia cases relating to novel Coronavirus (2019-nCOV). Told case incidence has been increasing dramatically and reached the hundreds, but it is likely to be an under-estimate[2]. The Coronavirus menace has now been spread all over the countries in globe. As of March 24, 2020, there were 372757 confirmed Novel Coronavirus-infected pneumonia (COVID-19) cases in 196 countries of which 16224 death cases were traced out worldwide[3]. This endemic of novel Coronavirus has been caused due to the mobility and interaction of people who moved from one country to another by outbound fights. Therefore, we argue that densely populated countries as well as cities are more likely to be infected with novel Coronavirus because of inbound and outbound flights and other means of transports. It implies that both in-bound and out-bound flights of passengers are significant factors in triggering Coronavirus infection risk in more densely populated countries than in less densely populated countries. However, previous studies did not consider the population density factor while modeling the risk of 2019-nCOV transmission. Recent studies estimates the risk of Coronavirus transmission from the Chinese cities to other destinations[4,5]. More importantly, Haider et al. (2020) built a risk index using the number of air travelers multiplied by the weight of the number of infected cases. Apart from previous studies, our study can fill the research gap constructing a risk index that takes population density and size, case identification rate, and the number of air travel into account.

## 2 Methods

### 2.1 Risk Index

We present a mathematical model that assesses the risk of infectious diseases such as Coronavirus. The model includes population size and density, the number of identified cases, case identification rate, and passengers’ travel among different regions. Risk Index (RI) of a certain infectious disease in a specific area *a*_*ij*_ of a country *C*_*i*_ where *i* = 1,2,3,…, *n* and *j* = 1,2,3, …, *m*_*i*_ is estimated for a given Calculation Period (CP) which is usually days, weeks, months or even years. RI is unit free which makes it comparable among areas; higher the RI, the greater the risk. Therefore, RI of an area *a*_*ij*_ is defined as

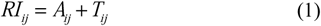

where, *A*_*ij*_ and *T*_*ij*_ are the area and the travel specific risk components respectively.

#### 2.1.1 Area specific risk

Area specific component estimates the risk of infectious diseases posed by the factors such as the number of identified cases, population size, and case identification rate. We also consider mobility—interaction factor to estimate the risk of spreading infectious diseases as mobility and interaction among people contribute to spread infectious diseases according to some studies [6–8]. So, it is expected that the higher the mobility and interaction among people in an area, the higher the risk of spreading infectious diseases. We can call it “*Mobility-Interaction Effect”*. The reasoning behind such hypothesis is that people have the tendency to amass in an area which provides better environment, livelihood, education, and other amenities. The aggregation process continues further with the economic development, forcing people to move low density areas to high density areas. As a result, people living in high density areas have higher mobility and interaction among themselves than the people living in low density areas[9]. Therefore, we propose that *Relative Mobility-Interaction Effect* (RMIE) can be estimated by the relative population density of that area within a country.

For an area *a*_*ij*_, having the population density *D*_*ij*_ in country *C*_*i*_, RMIE is measured as

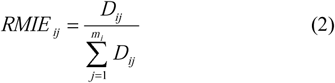

Now using RMIE along with the number of identified cases, case identification rate, calculation period of risk measurement, and assuming that infectious disease spread at geometric rate, we write the area specific risk component as

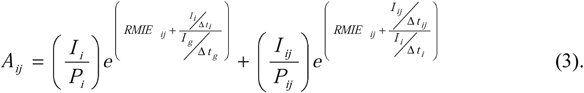

Here, within the risk calculation period (CP),

*P*_*i*_ = Population of country *i*

*P*_*ij*_ = Population of area/city *a*_*ij*_ of country *i*

*I* _*i*_ = The total identified case of infectious disease in country *i*

*I* _*ij*_ = The total identified cases of infectious disease in *a*_*ij*_

*t* _*s*_ = Starting time of CP

*t* _*e*_ = Ending time of CP

*t*_*ij*_ = Time when first case was identified in area/city *a*_*ij*_

*t* _*i*_ = Time when first case was identified in country *i*

*t* _*g*_= Time when first case was identified globally

Δ*t*_*g*_ = *t*_*e*_ − *t* _*g*_ the duration of global identification

Δ*t*_*i*_ = *t*_*e*_ − *t*_*i*_ the duration of first case identification in country *i*

Δ*t*_*ij*_ = *t*_*e*_ − *t* _*ij*_ the duration of first case identification in area *a*_*ij*_.

It accumulates two risk components: one is the area/city risk and the other is the country risk. An important feature of the measure (3) is that if an area has no identified case, we can assess area specific risk by country specific risk since through mobility-interaction effect of other areas create pressure of spreading disease to that particular area.

#### 2.1.2 Travel specific risk

Drawing experience from previous outbreak of infectious diseases including the recent epidemic of Coronavirus, literatures suggest addressing the risk of travelling among different areas and countries[10–12]. In our study, the travel specific risk component for area *a*_*ij*_ of country *C*_*i*_ is measure as

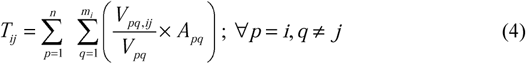

where *V*_*pq*_ is the number of travelers who left the area *a* _*pq*_ for different destinations within the time duration, *V*_*pq, ij*_ is the number of travelers who arrive at the area *a*_*ij*_ from *a*_*pq*_ within the same time duration, and *A*_*pq*_ is the area specific component for *a*_*pq*_ as described in equation 3. The travel risk component incorporates contribution of other area specific risks proportionately to the proportion of travelers from those areas. Combining equation (3) and (4), we get the final RI as follows:

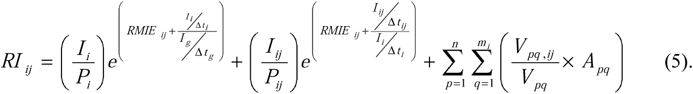

Some interesting features of this index are worthwhile to mention. The index says that there is a risk of Coronavirus spreading even though the travel specific risk component becomes zero. That means even if a country imposes air travel restriction internationally, the risk of transmission still exists. The area specific component of the index actually portrays the dynamics of infectious disease spreading influenced by mobility and interaction among people. One of the important implications of the area specific component is that restricting international movement is not sufficient to contain the risk of Coronavirus spreading in particular and infectious diseases in general. Internal restriction is also important to suppress transmission risk in an area which is exactly what the current situation says. In that sense, we expect that our index has the higher probability of mimicking the current situation of Coronavirus.

### 2.2 Data sources

Data on Coronavirus identified cases, population size and density, and air travel among areas of the four countries – US, Canada, Australia and China are collected from three different sources. First, the total number of confirmed cases (up to 26 March 2020) and the dates of first identification of Coronavirus cases in geographic areas (territory/province/state) are extracted from Johns Hopkins University Center for Systems Science and Engineering (JHU CCSE) (https://systems.jhu.edu/research/public-health/ncov/)[13,14]. Second, data on population size and geometric area are collected from the website https://www.citypopulation.de. Finally, because of insufficient passengers’ travel data, we incorporate air travel data only among these areas. Air travel data are obtained from FLIRT (https://flirt.eha.io), a flight network analysis tool developed by EcoHealth Alliance which provides simulated passengers’ data for the final destination from different airports based on flight schedule database of 800 airlines[12,15].

### 2.3 Data analysis

We have defined the calculation period (CP) from 31^st^ December 2019 to 27^th^ March 2020.The total number of confirmed Coronavirus cases are accumulated up to 26^th^ March 2020 for different areas of respective countries. Population density (population per square kilometer) of these areas is calculated by dividing population size by the respective geometric area. At first, we calculate area specific component using equation 3. To estimate travel specific risk component presented in equation 4, we calculate the proportion of travelers arrived at area *a*_*ij*_ from other areas directly by using simulated data from FLIRT. In total 781 international airports among these areas are included in this study. All analysis are performed using statistical software R version 3.6.0[16].

## 3. Results

In this study, we present Coronavirus risk ranking of 104 areas (territory/province/state) of four countries based on our developed risk index (RI). Table 1 presents the rank of these areas along with values of area specific component (A), travel specific component (T) and overall RI score as of time from 31 December 2019 to 26 March 2020.

**Table 1:** Areas based on risk index for Coronavirus transmission

| Rank | Area, Country (total cases) | A,T | RI | Rank | Area, Country (total cases) | A,T | RI |
| --- | --- | --- | --- | --- | --- | --- | --- |
| 1 | New York, US(38262) | 0.00678, 0.00014 | 0.00691484 | 53 | Nebraska, US(74) | 0.000363, 0.000036 | 0.00039865 |
| 2 | Hubei, China(70970) | 0.00291, 0.00002 | 0.00292621 | 54 | South Dakota, US(47) | 0.000378, 0.000011 | 0.00038858 |
| 3 | Massachusetts, US(2442) | 0.00072, 0.00148 | 0.00220135 | 55 | Victoria, Australia(523) | 0.000238, 0.000149 | 0.00038736 |
| 4 | District of Columbia, US(234) | 0.00112, 0.00087 | 0.00199245 | 56 | West Virginia, US(52) | 0.000354, 0.000011 | 0.0003649 |
| 5 | New Jersey, US(6957) | 0.0014, 0.00051 | 0.00190124 | 57 | Alberta, Canada(488) | 0.000287, 0.000074 | 0.00036152 |
| 6 | Georgia, US(1573) | 0.00049, 0.00086 | 0.00134612 | 58 | Shanghai, China(456) | 0.000119, 0.000233 | 0.00035187 |
| 7 | Colorado, US(1449) | 0.00059, 0.00058 | 0.00117628 | 59 | British Columbia, Canada(739) | 0.000302, 0.000044 | 0.00034597 |
| 8 | Illinois, US(2564) | 0.00054, 0.00063 | 0.00116654 | 60 | Beijing, China(574) | 0.000108, 0.00023 | 0.00033756 |
| 9 | Michigan, US(2906) | 0.00066, 0.0004 | 0.00106406 | 61 | Ontario, Canada(871) | 0.000216, 0.000115 | 0.00033119 |
| 10 | Maryland, US(587) | 0.00044, 0.00056 | 0.00099436 | 62 | Nova Scotia, Canada(73) | 0.000232, 0.000072 | 0.00030456 |
| 11 | Pennsylvania, US(1813) | 0.00048, 0.00048 | 0.0009651 | 63 | Newfoundland and Labrador, Canada(82) | 0.000288, 0.000013 | 0.00030029 |
| 12 | Utah, US(397) | 0.00045, 0.00051 | 0.00095981 | 64 | South Australia, Australia(235) | 0.000264, 0.000017 | 0.00028078 |
| 13 | Washington, US(3357) | 0.00079, 0.00016 | 0.00095008 | 65 | Queensland, Australia(494) | 0.000235, 0.000022 | 0.00025722 |
| 14 | Louisiana, US(2387) | 0.0009, 0.00005 | 0.00094985 | 66 | Western Australia, Australia(233) | 0.000223, 0.000015 | 0.00023805 |
| 15 | Minnesota, US(346) | 0.00039, 0.00045 | 0.00083765 | 67 | Tasmania, Australia(47) | 0.000213, 0.00001 | 0.00022362 |
| 16 | Nevada, US(430) | 0.00047, 0.00028 | 0.00074403 | 68 | Saskatchewan, Canada(95) | 0.000203, 0.00001 | 0.00021303 |
| 17 | Ohio, US(883) | 0.00041, 0.00033 | 0.00073724 | 69 | Tianjin, China(154) | 0.00009, 0.000105 | 0.00019453 |
| 18 | North Carolina, US(741) | 0.0004, 0.00033 | 0.00073243 | 70 | Prince Edward Island, Canada(5) | 0.000188, 0.000002 | 0.00019026 |
| 19 | Connecticut, US(1033) | 0.00065, 0.00005 | 0.0007 | 71 | Chongqing, China(584) | 0.000093, 0.000097 | 0.00019022 |
| 20 | Quebec, Canada(1640) | 0.00061, 0.00007 | 0.00068212 | 72 | Guangdong, China(1456) | 0.000088, 0.000102 | 0.00018973 |
| 21 | Florida, US(2386) | 0.00045, 0.00022 | 0.00067425 | 73 | Yukon, Canada(3) | 0.000186, 0.000003 | 0.00018931 |
| 22 | Texas, US(1584) | 0.00038, 0.00026 | 0.00064431 | 74 | Hainan, China(174) | 0.000092, 0.00009 | 0.00018223 |
| 23 | Arizona, US(516) | 0.0004, 0.00024 | 0.00063831 | 75 | New Brunswick, Canada(33) | 0.000171, 0.000006 | 0.00017732 |
| 24 | Vermont, US(167) | 0.00059, 0.00002 | 0.00061872 | 76 | Northern Territory, Australia(12) | 0.000163, 0.000007 | 0.00017001 |
| 25 | Tennessee, US(1100) | 0.0005, 0.00011 | 0.00060414 | 77 | Manitoba, Canada(36) | 0.00014, 0.000024 | 0.00016451 |
| 26 | California, US(3980) | 0.00043, 0.00017 | 0.00060279 | 78 | Liaoning, China(130) | 0.000076, 0.000087 | 0.00016307 |
| 27 | Missouri, US(529) | 0.00041, 0.00018 | 0.00059377 | 79 | Northwest Territories, Canada(1) | 0.000132, 0.00003 | 0.00016177 |
| 28 | Rhode Island, US(165) | 0.0005, 0.00005 | 0.00054924 | 80 | Zhejiang, China(1244) | 0.000097, 0.000062 | 0.00015922 |
| 29 | Australian Capital Territory, Australia(53) | 0.00053, 0.00002 | 0.00054771 | 81 | Henan, China(1297) | 0.000089, 0.00007 | 0.00015896 |
| 30 | Mississippi, US(491) | 0.00049, 0.00002 | 0.00051384 | 82 | Guizhou, China(148) | 0.000077, 0.00008 | 0.00015662 |
| 31 | Indiana, US(662) | 0.00043, 0.00007 | 0.0004952 | 83 | Shaanxi, China(256) | 0.000079, 0.000076 | 0.00015552 |
| 32 | Wisconsin, US(738) | 0.00045, 0.00004 | 0.00049232 | 84 | Fujian, China(329) | 0.000082, 0.000067 | 0.00014901 |
| 33 | South Carolina, US(433) | 0.00041, 0.00006 | 0.00047741 | 85 | Sichuan, China(550) | 0.000079, 0.000067 | 0.00014588 |
| 34 | Oregon, US(327) | 0.0004, 0.00007 | 0.00047166 | 86 | Nunavut, Canada(0) | 0.000109, 0.00001 | 0.00011991 |
| 35 | Delaware, US(131) | 0.00047, 0 | 0.00047103 | 87 | Heilongjiang, China(497) | 0.000085, 0.000033 | 0.00011853 |
| 36 | Kentucky, US(252) | 0.00038, 0.00009 | 0.00046989 | 88 | Anhui, China(996) | 0.00009, 0.000027 | 0.00011712 |
| 37 | Maine, US(155) | 0.00044, 0.00003 | 0.00046821 | 89 | Jervis Bay, Australia(0) | 0.000117, 0 | 0.00011678 |
| 38 | New Hampshire, US(138) | 0.00043, 0.00004 | 0.00046773 | 90 | Shandong, China(778) | 0.000083, 0.000031 | 0.00011446 |
| 39 | Arkansas, US(337) | 0.00044, 0.00002 | 0.00046019 | 91 | Guangxi, China(256) | 0.000078, 0.000035 | 0.00011316 |
| 40 | Alabama, US(518) | 0.00044, 0.00002 | 0.00045785 | 92 | Hunan, China(1022) | 0.000089, 0.000024 | 0.00011281 |
| 41 | Hawaii, US(95) | 0.0004, 0.00005 | 0.00044033 | 93 | Jiangsu, China(640) | 0.000084, 0.000026 | 0.00011039 |
| 42 | Oklahoma, US(255) | 0.00039, 0.00005 | 0.00043938 | 94 | Yunnan, China(180) | 0.000076, 0.000034 | 0.00010951 |
| 43 | Idaho, US(149) | 0.00041, 0.00002 | 0.00043153 | 95 | Qinghai, China(18) | 0.000075, 0.000032 | 0.0001067 |
| 44 | Wyoming, US(53) | 0.00042, 0.00001 | 0.00042825 | 96 | Jiangxi, China(937) | 0.000094, 0.000012 | 0.00010616 |
| 45 | Montana, US(90) | 0.00041, 0.00001 | 0.00042416 | 97 | Ningxia, China(75) | 0.000083, 0.000022 | 0.00010499 |
| 46 | New Mexico, US(114) | 0.00038, 0.00004 | 0.00042241 | 98 | Hebei, China(325) | 0.000078, 0.000026 | 0.00010426 |
| 47 | Virginia, US(476) | 0.00038, 0.00003 | 0.00041985 | 99 | Shanxi, China(135) | 0.000076, 0.000021 | 0.00009768 |
| 48 | Kansas, US(175) | 0.00039, 0.00003 | 0.00041882 | 100 | Xinjiang, China(79) | 0.000075, 0.00002 | 0.00009434 |
| 49 | New South Wales, Australia(1226) | 0.00037, 0.00005 | 0.00041665 | 101 | Jilin, China(96) | 0.000076, 0.000018 | 0.00009381 |
| 50 | Alaska, US(57) | 0.0004, 0.00001 | 0.00041182 | 102 | Inner Mongolia, China(90) | 0.000075, 0.000017 | 0.00009202 |
| 51 | Iowa, US(180) | 0.00038, 0.00003 | 0.00041118 | 103 | Tibet, China(1) | 0.000072, 0.000015 | 0.0000865 |
| 52 | North Dakota, US(51) | 0.00039, 0.00001 | 0.00040226 | 104 | Gansu, China(138) | 0.000077, 0.000003 | 0.00007995 |

**Figure 1:**
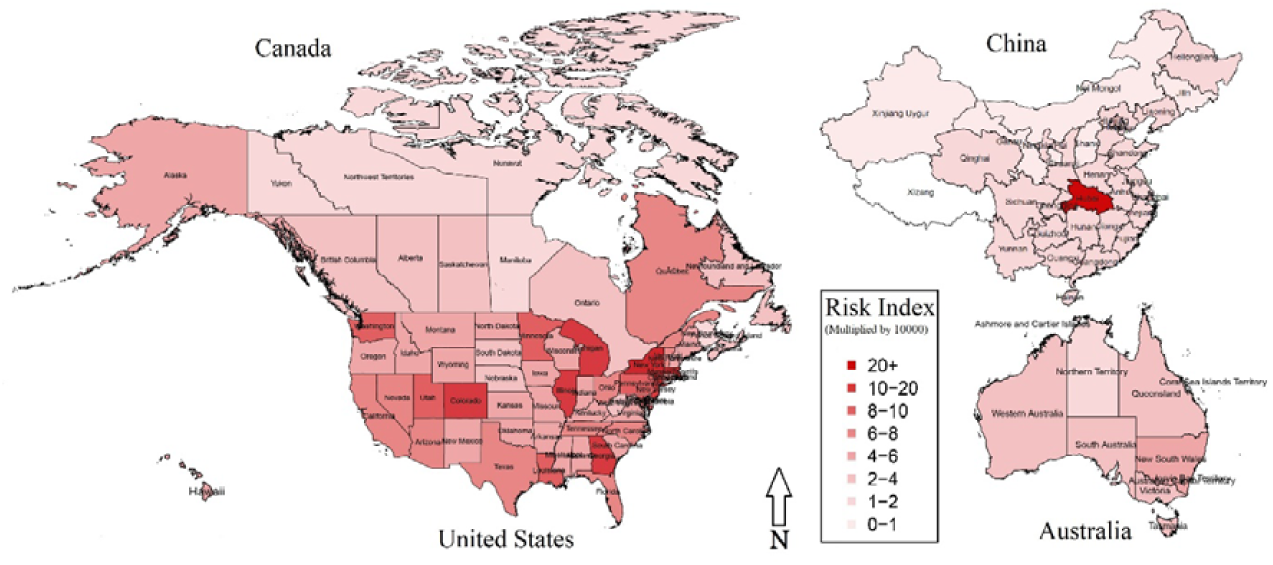
Risk Index of Coronavirus infection (as of 26th March 2020)

Empirical findings depict that New York State in US is the most risky area in terms of Coronavirus infection among US, Canada, Australia, and China. As per report of 26 March 2020, about 38262 Coronavirus cases are detected in this state, and the value of RI is 0.00691484 which clearly shows the severity of infection comparing to others. Besides, some other areas such as Hubei (China), Massachusetts (US), District of Columbia (US), New Jersey (US), Georgia (US), Colorado (US), Illinois (US), Michigan (US) and Maryland (US) are ranked 2nd, 3rd, 4th, 5th, 6th, 7th, 8th, 9th and 10th respectively as per calculation of RI. Besides, Quebec is ranked 20th whereas Alberta is ranked 57th among Provinces in Canada. Moreover, Australian Capital Territory, Australia is ranked 29th followed by New South Wales in 49th position. More interestingly, remaining Chinese Provinces are ranked at the bottom of the table, implying less risk of Coronavirus infection in China comparing to US, Canada and Australia.

## 4. Discussion

China (Hubei Province) as a breeding ground was the highest in the importation to Coronavirus infection across the globe. Later, this pandemic rapidity of this disease has now captured US and some other parts of the world. It has been spread at a geometric rate due to the mobility and interaction of people through air-borne movement from one country to another or from one city to another. In this respect, our study models a risk index to measure the human to human Coronavirus transmission mainly caused by people’s mobility and interaction. Therefore, we recommend avoiding any mass gathering, maintaining recommended physical distance and restricting inbound and outbound flights across different areas/cities and countries as well.

## Data Availability

All data are publicly available.
1. The total number of confirmed COVID-19 cases (up to 26 March 2020) and the dates of first identification in geographic areas (territory/province/state) are extracted from Johns Hopkins University Center for Systems Science and Engineering (JHU CCSE) (https://systems.jhu.edu/research/public-health/ncov/) Current Github repository - https://github.com/CSSEGISandData/COVID-19
2. Data on population size and geometric area are collected from the website https://www.citypopulation.de 
3. Air travel data are obtained from FLIRT (https://flirt.eha.io), a flight network analysis tool developed by EcoHealth Alliance.

## References

1 Sumdani H, Frickle S, Le M, et al. Effects of Population Density on the Spread of Disease.

2 Nishiura H, Jung S, Linton NM, et al. The Extent of Transmission of Novel Coronavirus in Wuhan, China, 2020. J Clin Med 2020;9:330. doi:10.3390/jcm9020330

3 World Health organization (WHO). Coronavirus disease 2019 (COVID-19) Situation Report – 64. 2020. https://www.who.int/docs/default-source/coronaviruse/situation-reports/20200324-sitrep-64-covid-19.pdf?sfvrsn=703b2c40_2

4 Chinazzi M, Davis JT, Ajelli M, et al. The effect of travel restrictions on the spread of the 2019 novel coronavirus (COVID-19) outbreak. Science (80-) 2020;:eaba9757. doi:10.1126/science.aba9757

5 Bogoch II, Watts A, Thomas-Bachli A, et al. Potential for global spread of a novel coronavirus from China. J Travel Med 2020;27. doi:10.1093/jtm/taaa011

6 Crooks AT, Hailegiorgis AB. An agent-based modeling approach applied to the spread of cholera. Environ Model Softw 2014;62:164–77. doi:10.1016/j.envsoft.2014.08.027

7 Wesolowski A, Eagle N, Tatem AJ, et al. Quantifying the Impact of Human Mobility on Malaria. Science (80-) 2012;338:267–70. doi:10.1126/science.1223467

8 Prothero RM. Disease and mobility: a neglected factor in epidemiology. Int J Epidemiol 1977;6:259–67.

9 Fujimoto S, Mizuno T, Ohnishi T, et al. Relationship between population density and population movement in inhabitable lands. Evol Institutional Econ Rev 2017;14:117–30.

10 Stoddard ST, Morrison AC, Vazquez-Prokopec GM, et al. The Role of Human Movement in the Transmission of Vector-Borne Pathogens. PLoS Negl Trop Dis 2009;3:e481. doi:10.1371/journal.pntd.0000481

11 Epstein JM, Goedecke DM, Yu F, et al. Controlling Pandemic Flu: The Value of International Air Travel Restrictions. PLoS One 2007;2:e401. doi:10.1371/journal.pone.0000401

12 Haider N, Yavlinsky A, Simons D, et al. Passengers’ destinations from China: low risk of Novel Coronavirus (2019-nCoV) transmission into Africa and South America. Epidemiol Infect 2020;148:e41. doi:10.1017/S0950268820000424

13 Dong E, Du H, Gardner L. An interactive web-based dashboard to track COVID-19 in real time. Lancet Infect Dis Published Online First: February 2020. doi:10.1016/S1473-3099(20)30120-1

14 JHU CSSE. 2019 Novel Coronavirus COVID-19 (2019-nCoV) Data Repository by Johns Hopkins CSSE. 2020.https://github.com/CSSEGISandData/COVID-19 (accessed 27 Mar 2020).

15 EcoHealth alliance. FLIRT: a product of EcoHealth alliance. 2020.https://flirt.eha.io (accessed 16 Mar 2020).

16 Team RC. R: a language and environment for statistical computing computer program. R Core Team, Vienna, Austria. 2019.

